# Trustworthy personalized treatment selection: causal effect-trees and calibration in perioperative medicine

**DOI:** 10.64898/2026.03.03.26347440

**Authors:** Yoel Mittelberg, Daniel K. Stiglitz, Gideon Kowadlo

## Abstract

**Background:** Personalized medicine promises to tailor treatments to the individual, but it carries a hidden risk: mistaking statistical noise for actionable clinical insight. Current machine learning approaches often provide predictions, but fail to inform clinicians when those predictions are unreliable.

**Objective:** Develop a deployment-readiness framework that integrates causal inference, interpretable effect-trees, and calibration assessment to distinguish actionable signal from unreliable variation, and to support treatment selection only when the estimated benefit is both reliable and clinically meaningful.

**Methods:** Using retrospective observational cohort EHR data from the INSPIRE perioperative dataset (N>130,000 surgical operations, 2011–2020), we estimated treatment effects using causal forests with double machine learning, benchmarked against other causal methods to assess convergence. We used the estimated causal effects to create effect-trees and translated estimates into interpretable rules. We validated the treatment recommendations by assessing subgroup calibration to identify which groups were reliable for treatment selection.

**Results:** In a prostate procedures case study (neuraxial versus general anesthesia; total N=2,822), neuraxial anesthesia was associated with substantially lower post-operative opioid use (ATE = −1.38 opioid medications, 95% CI [−1.62, −1.15]). The effect-tree produced five clinically interpretable subgroups using BMI, ASA status, and age, with effects ranging from −1.10 to −1.59 opioid medications. Calibration analysis identified four of five subgroups as reliable for deployment (calibration error < 0.08), while one small subgroup (N=250) showed higher calibration error (0.44), illustrating how the framework rates unreliable heterogeneity.

**Conclusions:** Individual prediction heterogeneity does not automatically justify clinical personalization. By combining effect-trees with calibration, this framework distinguishes actionable heterogeneity from noisy heterogeneity (detectable but unreliable). This approach transforms causal machine learning from a black box into a validated decision support system that enables selective deployment of treatment decision rules.

## 1 Introduction

Personalized medicine promises to tailor treatments to the individual, but it carries a hidden risk: mistaking statistical noise for actionable clinical insight. This can lead to treatment decision rules that are not effective or even harmful to the patient.

In perioperative medicine, clinicians routinely face decisions such as anesthesia selection, surgical approach and medication choices; these choices are often impacted by a large number of factors, with unique combinations of factors for each patient. Risk prediction models can be used to help make these decisions. However, systems that offer only risk predictions rather than causal effect estimates may fail to correctly guide these choices [1].

An alternative to risk prediction is causal inference, which aims to estimate the causal effect of a treatment on an outcome; for an individual patient, the difference in outcome between receiving and not receiving treatment. For example, given a specific medication (or exposure to a risk factor), is the patient more likely to experience a complication or not?

Observational EHR data offer unprecedented opportunities to study causal effects at scale, yet they are fraught with challenges due to the complex nature of confounding variables. Hernán and Robins [2] formalized a comprehensive framework for emulating clinical trials using real-world observational data. In observational studies, treatment assignment may not be random, requiring analysis to adjust for confounding in measured covariates as well as sensitivity analysis to gauge robustness to unmeasured confounding. In order to generate useful clinical insights, the challenge is threefold: we must adjust for confounding, estimate heterogeneity and crucially, translate complex black-box predictions into interpretable rules that clinicians can trust.

Heterogeneous treatment effect (HTE) estimation has become central to personalized medicine [1]. Modern causal inference methods, including meta-learners [3], causal forests with double machine learning [4, 5], and doubly robust estimators [6], enable individualized effect estimation. However, HTE analyses are only clinically useful when the variation in benefit and harm is large enough to span decision thresholds [7], and when predictions are sufficiently reliable to guide treatment decisions.

Calibration methods are used in risk prediction [8] and assess the reliability of model predictions by comparing predicted probabilities with observed event rates. In the context of causal inference, this is challenging, as individual-level treatment effects are never directly observable (each patient is either treated or not treated, but not both), but it can be done at a group level. Poorly calibrated models may incorrectly rank patients or grossly misestimate the magnitude of benefit, potentially leading to harmful over-treatment or missed opportunities for patient benefit. Recent work on causal inference has advanced calibration techniques [9] and heterogeneity testing methods [10], but frameworks for using calibration assessment to guide selective deployment decisions remain underdeveloped.

Policy trees partition patients using decision trees that take estimated treatment effects as inputs, creating human-readable treatment rules that clinicians can apply [11]. Recent approaches have expanded these methods by modifying the calculation of policy scores, implementing constraints and handling categorical and continuous variables [12] and emphasizing interpretability through two-step frameworks that separate CATE estimation from subgroup interpretation, using interpretable covariates (e.g., categorized age thresholds) for scientific communication in epidemiological research [13].

Despite advances in causal inference, few studies have translated heterogeneous treatment effect estimates into clinically interpretable treatment assignment rules, or established principled criteria for determining when those rules are sufficiently reliable for clinical use.

This study addresses both limitations by developing a **framework for interpretable and reliable selective deployment**, demonstrated on real-world EHR data from the INSPIRE perioperative dataset (N *>* 130,000 surgical operations). The framework combines causal effect estimation, effect-tree subgroup discovery, and calibration assessment. We adapt policy trees into effect-trees that explain variation in treatment response, translating individual treatment effect estimates into simple, clinician-readable subgroup rules. Calibration assessment then determines which subgroups are reliable enough for clinical use, operationalizing *selective deployment* : treatment recommendations are applied only where estimated benefits are well-calibrated and clinically meaningful. This approach moves causal machine learning from exploratory analysis to validated, selectively deployable decision support.

### 1.1 Paper organization

The remainder of this paper is organized as follows. Section 2 describes our methods, including the INSPIRE perioperative dataset, the data preprocessing pipeline, the causal inference framework, the construction of the effect-tree and calibration assessment. Section 3 presents results from the prostate opioids case study (neuraxial anesthesia effects on post-operative opioid use in prostate procedures). Section 4 discusses methodological contributions, implications for deployment readiness, study limitations, and future research directions. Supplementary materials provide population statistics, detailed results, and configuration settings.

## 2 Method

The main steps of the method are detailed in Figure 1.

**Figure 1:**
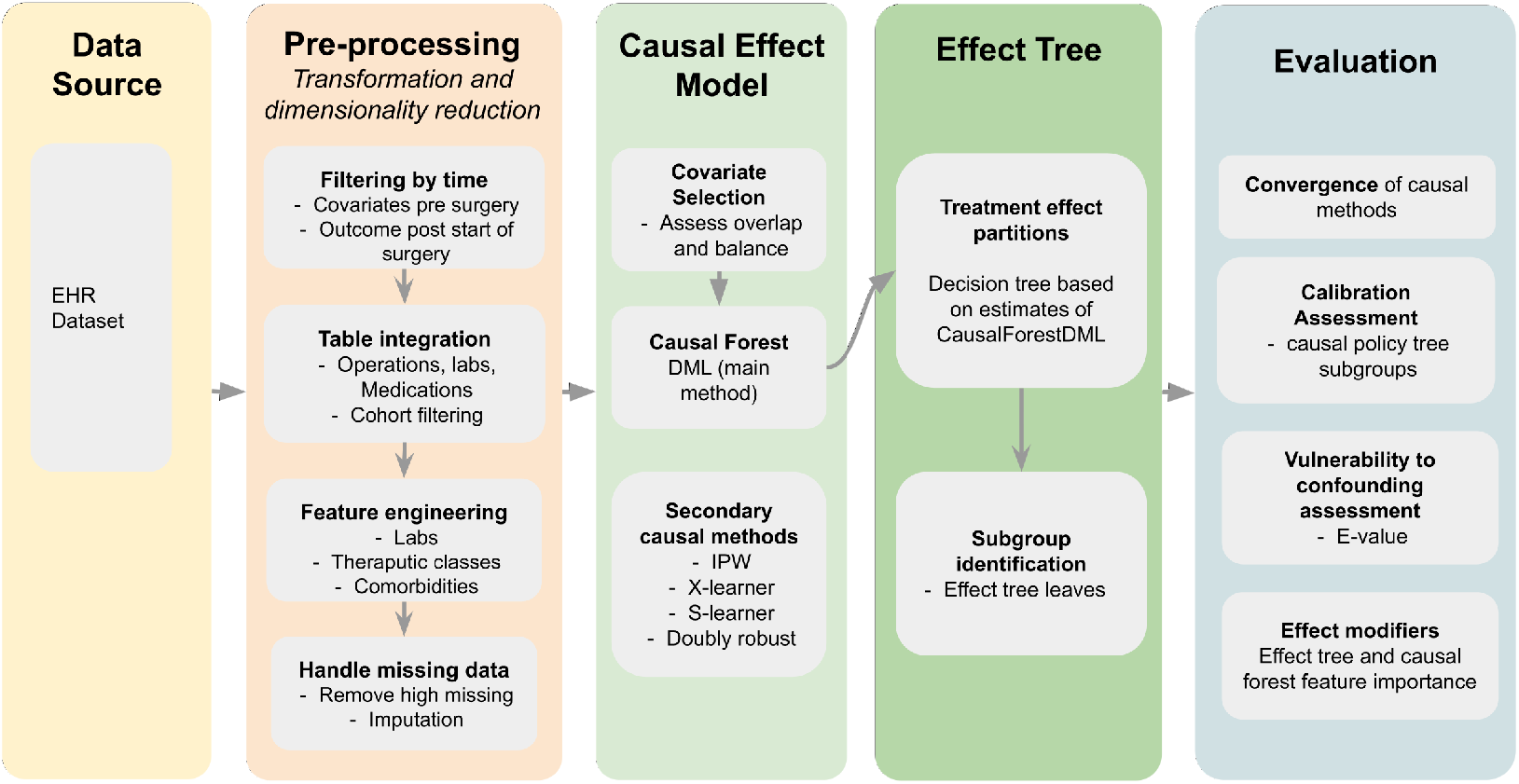
Key processing steps from data source to effect-tree and evaluation

### 2.1 Study design

We used a retrospective observational cohort design to evaluate treatment effects. The study used all eligible patients meeting the inclusion criteria for a case study selected on the basis of sufficient covariate balance, propensity score overlap, and a detectable treatment effect, conditions verified empirically prior to effect estimation.

#### Temporal ordering

Baseline features were measured before the start of surgery to ensure valid causal inference (up to 12 months before surgery).

#### No interference

We operated under the standard assumption that one patient’s treatment does not affect another’s outcome. The results section contains the cohort selection criteria defining the patient population, treatment definitions specifying exposure groups, and outcome definitions with measurement timeframes.

### 2.2 Data source

This study analyzed data from INSPIRE (INformed Surgical Patient care through Integrated REcord system) [14], a publicly available perioperative dataset from Seoul National University Hospital, South Korea. The dataset contains comprehensive EHR data for over 130,000 surgical operations of approximately 100,000 patients, covering the period January 2011 to December 2020 across multiple surgical specialties. INSPIRE includes detailed information about surgical procedures, pre- and post-operative laboratory values, medication administrations, and diagnoses coded in ICD-10-CM format. All data are de-identified to protect patient privacy.

Two data sources were excluded from the analysis. Vital signs were not included in order to focus on laboratory and clinical variables that are more standardized and measured consistently across diverse surgical contexts. Diagnoses were excluded because timestamps appear to indicate the time the diagnosis was recorded rather than the time it was made.

### 2.3 Data preprocessing

Raw EHR data from INSPIRE were preprocessed through a systematic pipeline consisting of 4 stages:

1. Filtering by time: raw data tables were filtered to prevent post treatment information from being captured by covariates.
2. Data integration: data were integrated across surgical operations, laboratory tests and medications and cohort filtering by demographics, procedure type, or other variables. Sparsity filtering was applied (*>*95% missing pre-aggregation, *>*70% post-aggregation, with exceptions for medication and laboratory indicators expected to be sparse). Outliers were detected using clinical reference ranges and duplicate records were removed.
3. Feature engineering: created treatment/outcome variables, derived clinical indicators (BMI, diagnostic categories) and therapeutic class exposures.
4. Missing data imputation: continuous and categorical variables were imputed using median and mode imputation for continuous and categorical variables, respectively. Missing medication values were treated as absent.

Covariates were entered on their natural scales after preprocessing and imputation, with no additional manual transformations.

### 2.4 Causal analysis framework

Our goal was to distinguish true treatment effects from spurious correlations driven by confounding. We organized variables into three roles:

- Treatment: the intervention whose effect we estimate.
- Outcome: the clinical endpoint measured after the start of surgery.
- Covariates: pre-treatment characteristics requiring adjustment.

Hypothesis-based covariates were created (as part of feature engineering) by using the following:

- Patient demographics
- The first 3 characters of ATC medication code as therapeutic class grouping and counting the occurrence of each group
- Counts of lab type occurrence
- The last value of each lab type prior to surgery
- Combinations of medications and labs as comorbidity indicators (e.g. diabetes)

### 2.5 Propensity score estimation

We used propensity scores (the probability of receiving treatment given a patient’s characteristics [15]) for inverse probability weighting to simulate the balance of a randomized trial, as well as for overlap assessment. Propensity scores were calculated using gradient boosting machine learning methods (XGBoost) to capture complex non-linear treatment selection patterns and interactions between covariates. The models used 100 trees with a maximum depth of 4, with 5-fold cross-validation.

### 2.6 Assessing causal inference validity

The adequacy of covariate characteristics to enable causal inference was evaluated through:

- Covariate balance assessment using standardized mean differences, with SMD *<* 0.1 considered good, and SMD *<* 0.25 considered acceptable (SMD *<* 0.1 has been proposed as indicating negligible imbalance [16]).
- Overlap examination of propensity score distributions to check positivity.

### 2.7 Causal effect estimation

We estimated the average treatment effect (ATE), the population-average outcome difference between treatment and control, and conditional average treatment effects (CATE) to identify subgroups with differential treatment benefit (heterogeneous treatment effects, HTE). We estimated these using five complementary methods and assessed their agreement as a robustness check. The five models were:

1. Causal forest double machine learning (CausalForestDML, econML) [4, 5]: Random forests to estimate individualized treatment effects, controlling for confounders through cross-validated residualization (DML) with honest sample splitting for valid inference. This was our primary method for heterogeneity estimation.
2. Inverse probability weighting (IPW) [2]: reweights observations by inverse propensity scores to create a pseudo-population where treatment is independent of confounders. We trimmed weights at the 0.10 quantile to limit instability. IPW was included for its simplicity and transparent confounder adjustment.
3. Doubly robust (DR) estimation [6]: combines propensity weighting and outcome modeling using gradient boosting and random forests. Only one of the models needs to be correctly specified for unbiased estimates. DR was included for robustness against model misspecification.
4. Meta-learners [3]: These use flexible machine learning models to estimate treatment effects. S-learner includes treatment as a covariate in a single outcome model. X-learner builds separate models for the treated and control groups and then imputes counterfactuals, making it particularly effective when treatment groups are imbalanced.

### 2.8 Using group effect-trees to identify causal effect subgroups

We used decision tree-based methods to extract interpretable clinical decision rules from individualized estimates, adapting concepts from the policy learning framework of Athey and Wager [11]. In standard policy learning, decision trees partition patients to maximize treatment benefit. We adapted this approach to focus on heterogeneity explanation rather than treatment optimization, creating what we term group effect-trees (effect-trees). Unlike black-box models, the resulting trees produce transparent subgroups that clinicians can identify using easily measured patient characteristics (e.g., age *>* 50 and BMI *<* 30).

Effect-trees were implemented using SingleTreeCateInterpreter from the EconML library. The algorithm takes individual-level CATE predictions from the causal forest DML model and patient covariates, then constructs a decision tree by maximizing variance in mean effect estimates across subgroups.

The following parameters of the tree interpreter have a significant impact on the identified subgroups:

- The maximum tree depth: the maximum number of levels of splitting in the tree (impacts the number of rules generated).
- The minimum leaf size: the minimum number of patients in a leaf of the tree (impacts the size of subgroup).
- The minimum impurity decrease: the minimum decrease in impurity required to split a node, where impurity is the variance of predicted treatment effects within a node (impacts the heterogeneity between subgroups).

These parameters are subjective and reflect a trade-off between fine-grained heterogeneity and estimate reliability. They should be set based on the objectives of analysis, and the resulting sub-groups assessed using calibration. Our objective was to generate a simple set of rules while identifying reliable heterogeneity. Therefore, we used a small maximum tree depth (3), a small but sufficient minimum leaf size to assess calibration (200), and unbounded minimum impurity decrease (the default value of 0). Estimator and analysis settings used in the case study are documented in Supplementary Table S5.

#### 2.8.1 Calibration assessment of heterogeneous treatment effects

Validating the calibration of predicted heterogeneous treatment effects is essential to ensure clinical reliability of targeted treatment recommendations. Calibration of standard predictive models assesses whether predicted probabilities match observed frequencies [8, 17]. We extend this concept to causal models, where calibration requires assessing whether predicted treatment effects match observed effects [9]. This cannot be assessed at the individual level, since each patient is observed under only one treatment condition, but it can be assessed at the group level.

We assessed calibration by comparing predicted conditional average treatment effects (CATE) with observed average treatment effects (ATE) within subgroups of patients. Calibration was evaluated across three types of subgroups to comprehensively assess model reliability: (1) effect-tree leaves, the decision rule subgroups produced by the effect-tree algorithm; (2) CATE quantiles, patients grouped by predicted treatment effect magnitude following the GATES methodology [10]; and (3) feature-based subgroups, defined by clinically meaningful characteristics such as age, sex, BMI, comorbidities, or disease severity (ASA).

For each subgroup *g*, calibration error was defined as:

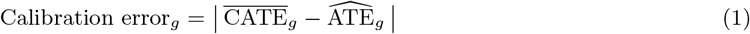

where 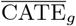 is the mean of individual treatment effect predictions within the subgroup, and 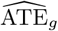 is the empirical difference in mean outcomes between treated and control patients in that subgroup. Errors are normalized by outcome standard deviation to allow assessment relative to outcome variability.

Calibration was used as a diagnostic tool to identify subgroups where predicted treatment effects are trustworthy versus where they require caution. We describe calibration as *good* when relative error is below 10% of outcome SD, *moderate* for 10–25%, and *poor* above 25%. These bands are heuristic, study-specific guides rather than universal clinical standards.

### 2.9 Implementation readiness assessment

Each subgroup was classified into one of three implementation readiness tiers using calibration bands of 10% (good) and 25% (moderate) of outcome SD, combined with effect magnitude and directional agreement between predicted and observed effects. Two effect magnitude thresholds were defined: a *meaningful* threshold for clinically significant outcomes (1 opioid medication count in this study), and a *minimal* threshold below which treatment effects are considered insufficient to justify implementation (0.2 medication counts in this study). These parameters are subjective and should reflect the clinical objectives of the specific analysis.

#### Implement

Good calibration (error *<* 10% of outcome SD), both predicted and observed effect magnitudes at or above the meaningful threshold, and directional agreement between predicted and observed effects.

#### Consider

Calibration is not classified as good, or effect magnitudes fall between the minimal and meaningful thresholds.

#### Do not implement

Predicted and observed effects disagree in direction, or either effect magnitude falls below the minimal threshold.

### 2.10 Evaluation

#### 2.10.1 Comparison of effect-tree drivers with other methods

Effect modifiers are variables that alter the causal effect of a treatment; unlike prognostic factors (which predict outcome regardless of treatment), they identify which patient characteristics indicate differential response to treatment. To assess whether effect-tree rules reflect causal drivers, we compared the variables used to split the effect-tree nodes with variables identified by other methods. If the variables used by effect-trees overlap those used by other methods, it increases confidence that they are genuine effect modifiers. We used two approaches for this assessment:

- Feature-based heterogeneity tests for clinically meaningful characteristics (age, ASA status, BMI, comorbidities): Spearman rank correlation between individual CATE estimates and each continuous or ordinal covariate, and Mann-Whitney U tests for binary covariates, to assess whether predicted treatment effects vary monotonically or systematically with each characteristic
- Causal forest feature importance: the relative contribution of each covariate to explaining variance in individual CATE estimates, providing a model-derived ranking of effect modifiers

#### 2.10.2 Sensitivity analysis

Observational studies face potential unmeasured confounding despite covariate adjustment. We quantified robustness through: (1) E-values [18] assessing the minimum confounder strength needed to negate observed effects, and (2) placebo tests on pre-treatment characteristics to detect residual confounding.

In addition, we compared the estimate from Causal Forest DML with other methods (IPW, doubly robust, meta-learners). Convergence of methods with different assumptions strengthens causal conclusions.

## 3 Results

### 3.1 Case study: neuraxial anesthesia and post-operative opioid use

We examined whether neuraxial anesthesia reduces post-operative opioid usage compared to general anesthesia in prostate procedure patients. The cohort comprised 2,822 male patients (aged 18–90) undergoing elective prostate procedures from INSPIRE, restricted to patients who received either general or neuraxial anesthesia. Prostate procedures were defined using ICD10-PCS codes, as procedure codes starting with “0VT0” or “0VB0” (prostate resection or excision). The stepwise cohort derivation is shown in Supplementary Figure S1 (Supplementary Section S1).

#### Treatment definition

Treatment was defined as receiving neuraxial anesthesia (*T* = 1) versus general anesthesia (*T* = 0). The cohort was restricted to these two anesthesia types; patients receiving other anesthesia types were excluded.

#### Outcome definition

The outcome variable was the post-operative opioid medication count within 48 hours of surgery (N02A ATC code medications).

#### Cohort

Of the 2,822 patients, 809 (28.7%) received neuraxial anesthesia and 2,013 (71.3%) received general anesthesia. We adjusted for four baseline covariates (used both for confounding adjustment and heterogeneity estimation): age, ASA physical status, BMI, and diabetes indicator. Prior opioid use was included in the configured adjustment set but was removed during analysis as a constant feature; all values were zero in this cohort, providing no discriminatory information.

### 3.2 Propensity score overlap and covariate balance

Covariate balance assessment revealed broadly acceptable balance between treatment groups (mean absolute SMD = 0.108; maximum absolute SMD = 0.236). ASA status (SMD = *−*0.061) and diabetes indicator (SMD = *−*0.003) met the *<* 0.1 threshold, while BMI (SMD = *−*0.131) showed moderate imbalance. Age (SMD = 0.236) showed the largest imbalance, approaching the 0.25 acceptability limit and warranting caution in interpreting age-related subgroup effects. Full baseline characteristics by treatment group and pre-imputation variable missingness are reported in Supplementary Tables S1 and S2, respectively.

Propensity score analysis revealed adequate overlap in treatment assignment (mean propensity score = 0.287, range: 0.004–0.973). Figure 2 shows overlap between treatment groups across the propensity score range, supporting causal conclusions.

**Figure 2:**
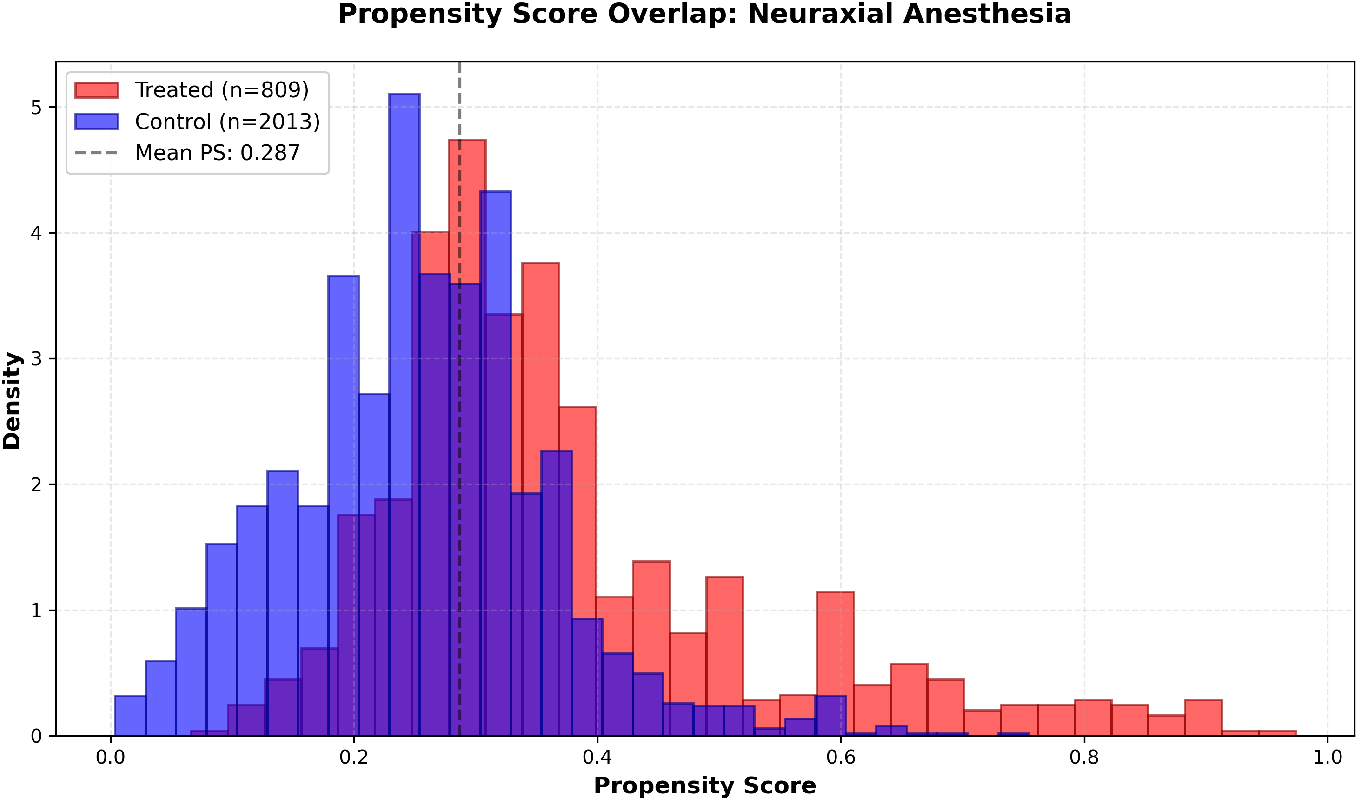
Propensity score distributions showing reasonable overlap between treated and control groups. Both treatment groups show broad distributions across propensity scores, indicating adequate common support for reliable causal inference.

### 3.3 Causal effect estimation

#### 3.3.1 Convergent estimates across methods

The average effect of treatment was estimated using five methods, which consistently estimated a reduction in opioid use, with covergent estimates between approaches, Table 1.

**Table 1:** Average treatment effect estimates across methods.

| Table 1: Average treatment effect estimates across methods |  |
| --- | --- |
| Method | ATE (Opioid medications) |
| Causal forest DML | $-1.383$ |
| Inverse probability weighting | $-1.338$ |
| Doubly robust | $-1.357$ |
| S-learner | $-1.312$ |
| X-learner | $-1.344$ |

The causal forest DML model estimated 1.38 fewer opioid medications (95% CI: [*−*1.62, *−*1.15]). The confidence interval excludes zero, and all five methods converge to similar estimates (range: *−*1.31 to *−*1.38), consistent with a protective effect of neuraxial anesthesia on post-operative opioid consumption.

#### 3.3.2 Heterogeneous treatment effects

The causal forest revealed meaningful heterogeneity in treatment effects (SD = 0.16, range: *−*1.70 to *−*1.06 opioid medications), with a median effect of *−*1.36 medications (Figure 3). All predicted individual effects were negative, suggesting consistent benefit across the cohort, though the magnitude varies substantially. This variation motivates the effect-tree analysis to identify which patient characteristics explain the differential magnitude of response and whether the resulting subgroup rules are reliable enough for clinical use.

**Figure 3:**
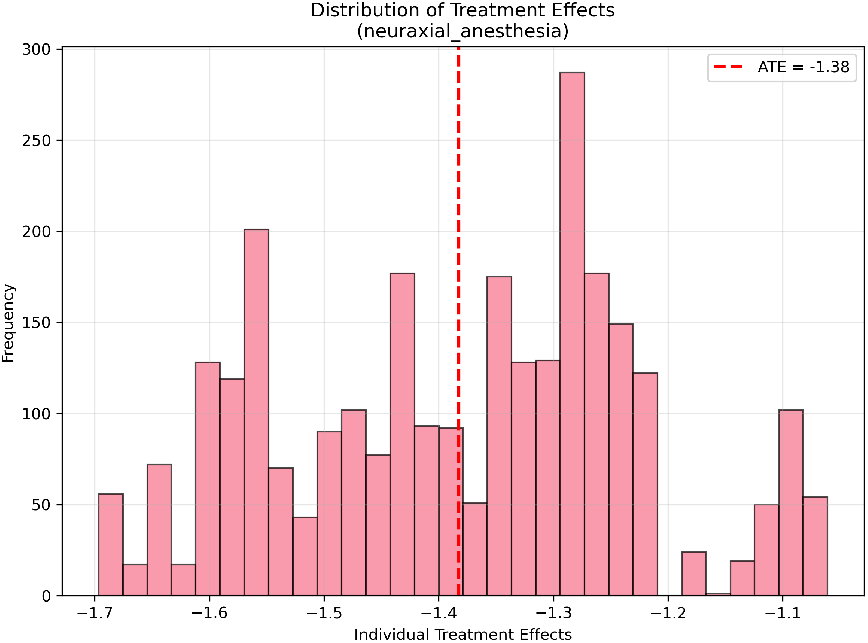
Heterogeneous treatment effect distribution (SD = 0.16 opioid medications)

### 3.4 Group effect-tree: interpretable treatment decision rules

The effect-tree (max depth 3, minimum leaf size 200, minimum impurity decrease 0) produced five subgroups using three clinical variables: BMI, ASA physical status, and age. The subgroup effects are shown in Figure 4 and summarized in Table 2, including each group’s predicted CATE, observed ATE, and calibration error.

**Table 2:** Effect-tree treatment decision rules (ordered by predicted CATE)

| Group | Decision rule | N | CATE | Obs. ATE | Cal. error |
| --- | --- | --- | --- | --- | --- |
| 1 | $\text{BMI} \leq 22.87 \ \& \ \text{ASA} \leq 1.5$ | 250 | $-1.10$ | $-0.66$ | 0.44 |
| 2 | $\text{BMI} \leq 22.87 \ \& \ \text{ASA} > 1.5$ | 592 | $-1.29$ | $-1.35$ | 0.05 |
| 3 | $\text{BMI} > 22.87 \ \& \ \text{age} \leq 72.5 \ \& \ \text{BMI} \leq 26.28$ | 993 | $-1.34$ | $-1.27$ | 0.07 |
| 4 | $\text{BMI} > 22.87 \ \& \ \text{age} \leq 72.5 \ \& \ \text{BMI} > 26.28$ | 468 | $-1.49$ | $-1.47$ | 0.02 |
| 5 | $\text{BMI} > 22.87 \ \& \ \text{age} > 72.5$ | 519 | $-1.59$ | $-1.56$ | 0.04 |

**Figure 4:**
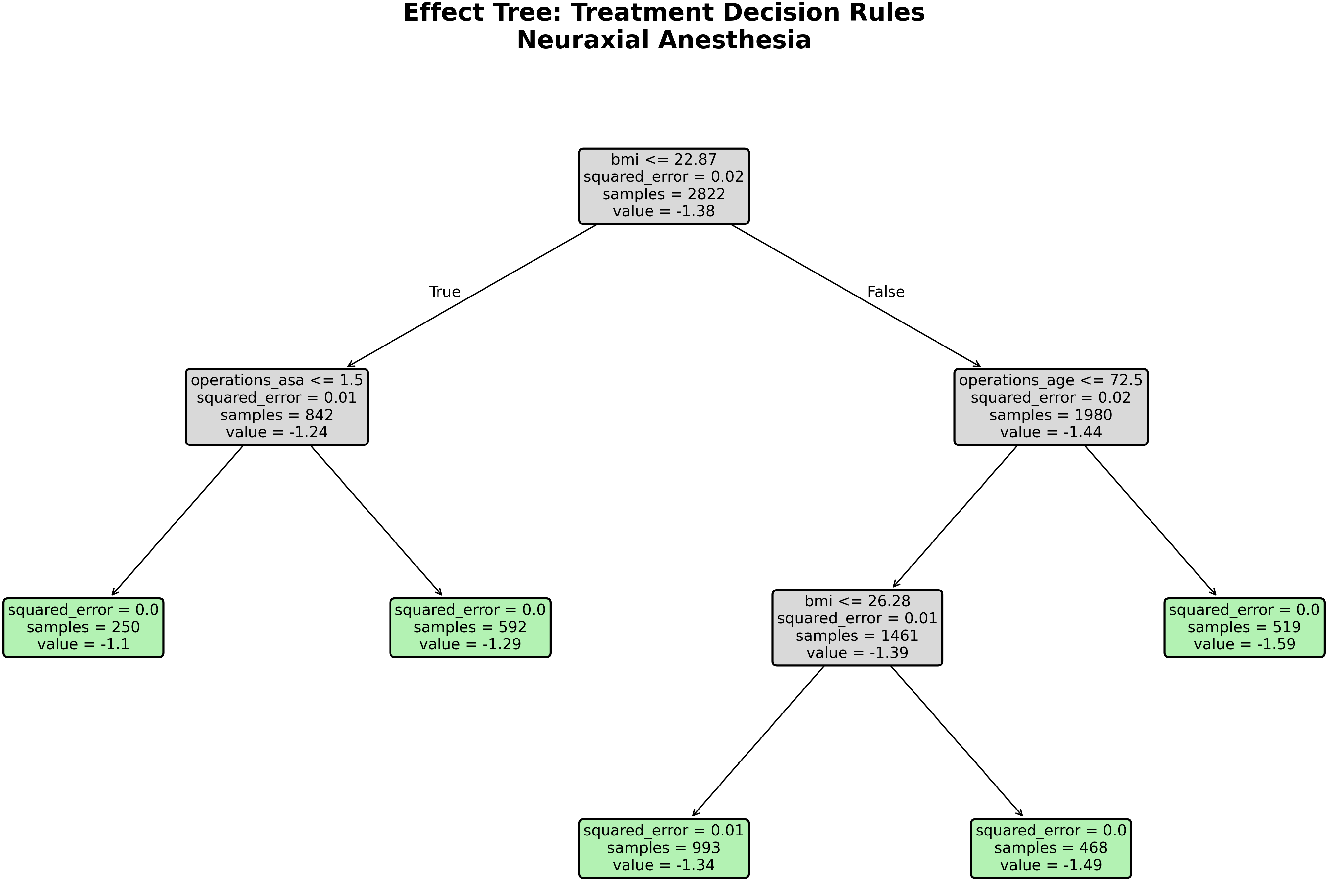
Effect-tree subgroup effects. The tree identified five subgroups using BMI, ASA status, and age, with predicted treatment effects (CATE) ranging from −1.10 to −1.59 opioid medications. Error bars show model-based confidence intervals.

#### Key clinical insights

The primary split was on BMI (threshold 22.87), separating lower-BMI patients (Groups 1, 2) from higher-BMI patients (Groups 3 – 5). Among lower-BMI patients, ASA status was the key differentiator: healthier patients (Group 1, ASA *≤* 1.5) showed the smallest predicted benefit (CATE = *−*1.10), while those with higher comorbidity burden (Group 2, ASA *>* 1.5) showed greater benefit (CATE = *−*1.29). Among higher-BMI patients, older age (*>*72.5 years, Group 5) was associated with the largest benefit (CATE = *−*1.59), while younger patients were further differentiated by BMI (Groups 3, 4). The overall pattern suggests that higher BMI and older age may be markers of greater opioid-reducing benefit from neuraxial anesthesia,

#### Calibration

Across predicted-effect quintiles (GATES), the causal forest DML reliably ranked patients by benefit magnitude (mean calibration error 0.11 opioid medications, 6.9% of outcome SD = 1.62; predicted vs. observed correlation *r* = 0.937; detailed results in Supplementary Table S3). At the subgroup level, calibration was good for Groups 2–5 (errors 0.02–0.07), but poor for Group 1 (error 0.44; predicted *−*1.10, observed *−*0.66). Group 1 comprises low-BMI, low-ASA patients (N=250, the smallest subgroup with only 82 treated patients), where the causal forest DML overestimates the magnitude of benefit. This illustrates the value of calibration assessment: a treatment rule based on predicted CATE alone would not have identified this subgroup as unreliable.

### 3.5 Implementation readiness assessment

Using calibration bands of *±*10% (good, *<* 0.16 opioid medications) and *±*25% (moderate, *<* 0.40) of outcome SD (= 1.62), a meaningful effect threshold of 1 opioid medication, and a minimal threshold of 0.2 medications (Figure 5), subgroups were classified as follows:

**Figure 5:**
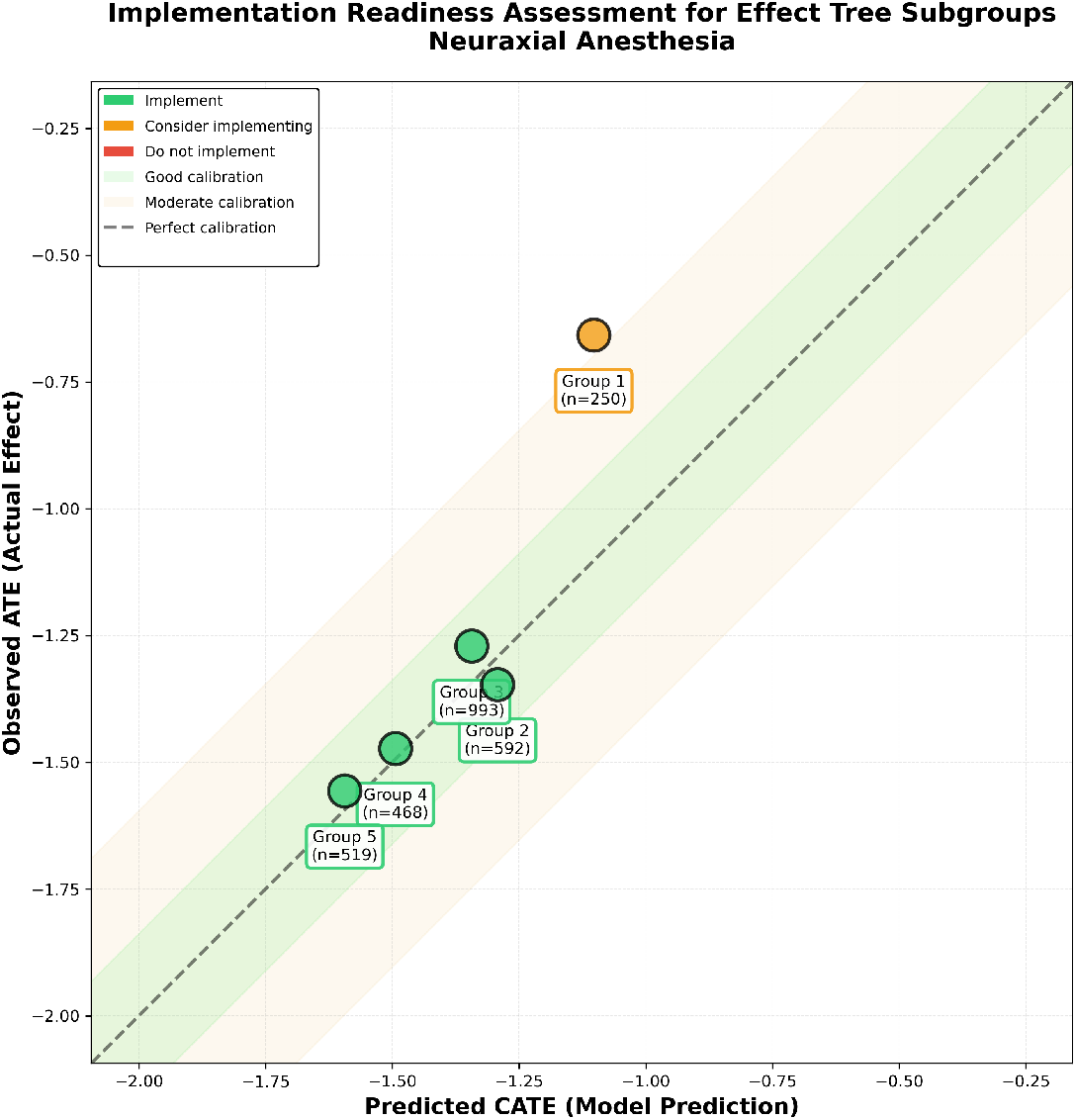
Implementation readiness assessment for effect-tree subgroups. Points near the diagonal indicate good calibration. Shaded bands represent good (±10% of outcome SD) and moderate (±25% of outcome SD) calibration regions. Groups 2 - 5 cluster near the diagonal; Group 1 deviates substantially.

- **Implement** (Groups 2 – 5): Four subgroups (N=2,572; 91.1% of cohort) show large effect magnitudes (*−*1.29 to *−*1.59 medications) with good calibration (errors 0.02 - 0.07). These subgroups support deployment of neuraxial anesthesia recommendations.
- **Consider** (Group 1): One subgroup (N=250; 8.9%) shows a moderate effect (*−*1.10 medications) but with high calibration error (0.44). The observed benefit (*−*0.66) is substantially smaller than predicted, warranting caution and further investigation before deploying targeted recommendations for this sub-group.

### 3.6 Evaluation

BMI, age, and ASA status showed evidence of effect modification in this cohort, consistent with the effect-tree using all three variables for subgroup identification.

#### 3.6.1 Feature importance: treatment assignment vs. effect modifiers

Distinguishing between variables that drive treatment selection (potential confounders) versus those that moderate treatment effects (effect modifiers) is critical for targeted medicine. The strongest treatment assignment predictors and effect modifiers were identified through propensity model and causal forest DML feature importance, respectively (Figure 6).

**Figure 6:**
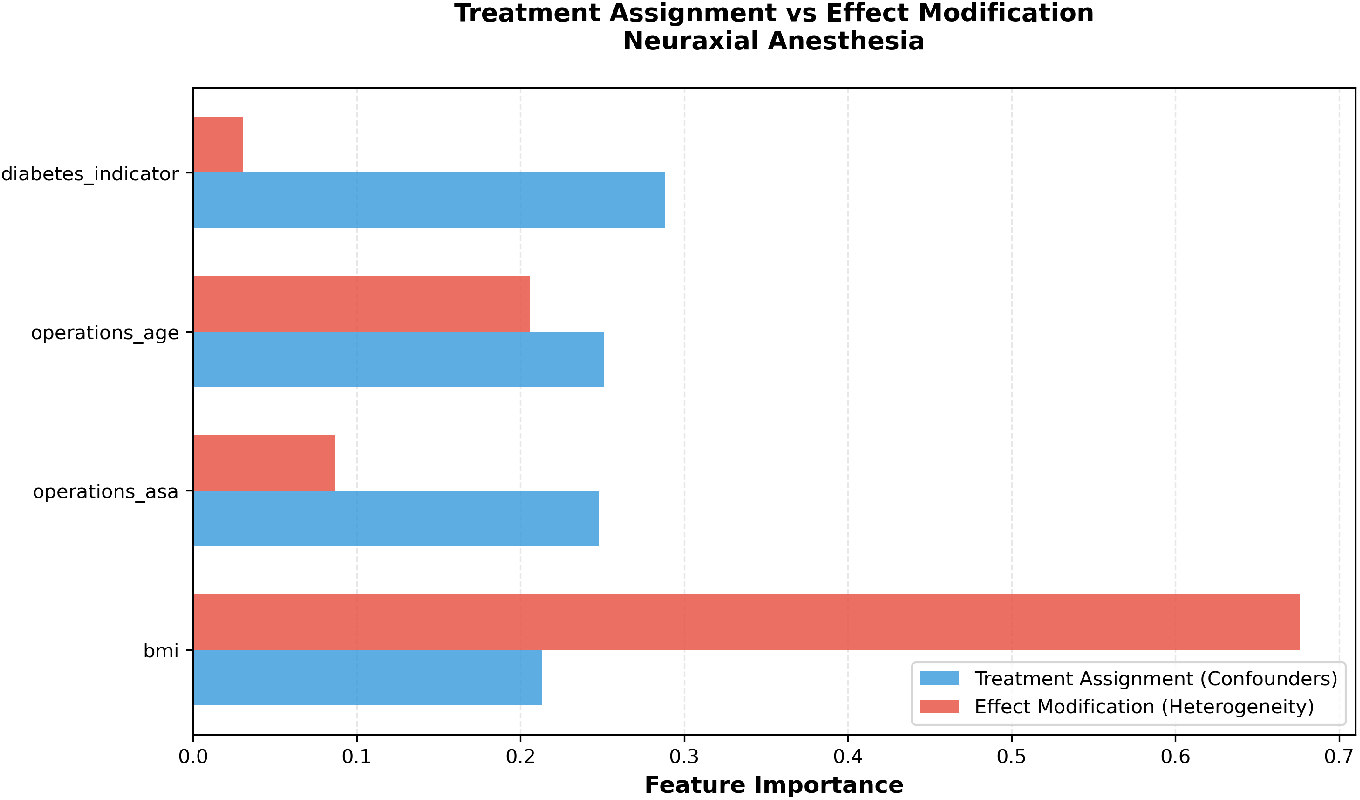
Feature importance: propensity model vs causal forest. BMI and age emerged as the strongest moderators in the causal forest, while propensity feature importance highlighted all variables as having similar importance, with diabetes the highest predictor of treatment assignment.

BMI was the dominant effect modifier (causal forest DML feature importance: 0.676), followed by age (0.206) and ASA status (0.087), with diabetes contributing minimally (0.031). These findings align with the effect-tree, which used BMI as the primary splitting variable and age and ASA as secondary splits. Feature-based heterogeneity tests provide a third source of convergence: Spearman correlations between each covariate and individual CATE estimates followed the same rank order as causal forest feature importance: BMI (*r* = *−*0.588), age (*r* = *−*0.385), ASA status (*r* = *−*0.229), confirming that patients who rank higher on these covariates consistently rank higher on predicted benefit. Statistical results are provided in Supplementary Table S4; individual CATE estimates plotted against each covariate are shown in Supplementary Figure S2. The propensity model identified a different pattern: diabetes was the strongest predictor of treatment assignment, while BMI and age had lower propensity importance. This distinction matters – variables that drive treatment selection are not necessarily the same as those that moderate treatment response.

#### 3.6.2 Sensitivity analysis

We employed sensitivity analyses to assess robustness to unmeasured confounding. E-value analysis (E= 3.78, CI lower bound 3.23) indicated high robustness: an unmeasured confounder would need to be associated with both treatment and outcome by a risk ratio of at least 3.78 to explain away the observed effect. Placebo tests showed no systematic bias (placebo treatment: p= 0.90; dummy outcome: p= 0.90).

The directional consistency across all five estimation methods — each with different assumptions (IPW relies on propensity scores, doubly robust protects against model misspecification, S-learner and X-learner use flexible machine learning, causal forest captures heterogeneity) provides reassurance that the observed effect is not an artifact of any single modeling choice. Together, the convergent estimates and passed placebo tests are consistent with a genuine protective effect of neuraxial anesthesia on post-operative opioid consumption.

## 4 Discussion

As healthcare systems race to implement AI-driven decision support, the distinction between detectable statistical patterns and reliable clinical rules becomes paramount. Our study leverages causal inference to estimate patient benefit (as opposed to calculating risk) and proposes a deployment-readiness workflow that couples interpretable effect-trees with calibration and effect-size assessment.

### 4.1 Principal findings

Our analysis demonstrates both the promise and practical challenges of data-driven targeted treatment strategies in perioperative medicine. In 2,822 prostate procedure patients, neuraxial anesthesia was associated with substantially reduced opioid use (ATE = *−*1.38 medications, 95% CI [*−*1.62, *−*1.15]), with high convergence across five estimation methods (range: *−*1.31 to *−*1.38). The effect-tree identified five clinically interpretable subgroups using BMI, ASA status, and age, with four of five groups (91.1% of patients) showing both large effects and good calibration. The fifth group, low-BMI, low-ASA patients, demonstrated high calibration error despite a predicted benefit, illustrating how the framework can identify subgroups where predicted effects should not be trusted for clinical decision-making.

### 4.2 Contributions

#### 4.2.1 Effect-trees as an interpretability layer

Our effect-trees build on the policy tree framework of Athey and Wager [11], adapting their approach from welfare optimization to heterogeneity explanation.

In the prostate opioids study, the effect-tree produced five subgroups using three clinical variables. The resulting rules are transparent and clinically interpretable: rather than presenting a clinician with raw predictions like *this patient’s predicted effect is −*1.34 *opioid medications*, decision trees provide actionable guidance (e.g., *for patients with BMI > 22*.*87, age ≤ 72*.*5, neuraxial anesthesia reduces opioid use by approximately 1*.*3–1*.*5 medications*), which clinicians can evaluate against their clinical knowledge. These quantified estimates may themselves be novel: while clinicians may know that neuraxial anesthesia tends to reduce opioid use, knowing that the benefit is approximately 1.3–1.5 medications, not just directionally positive, transforms qualitative intuition into an explicit magnitude that can be weighed in clinical decisions. Furthermore, the magnitude of subgroup effects enables explicit clinical trade-off reasoning: even when all patients are predicted to benefit, a larger predicted effect (e.g., Groups 4–5, CATE *−*1.49 to *−*1.59) may support a stronger recommendation when competing considerations such as contraindications or patient preference are present, while a smaller predicted effect (e.g., Group 1, CATE *−*1.10) may tip the decision differently depending on individual circumstances.

#### 4.2.2 Personalization granularity vs. calibration reliability

Effect-trees act as a safeguard against spurious personalization: splitting stops when no further partition would meaningfully separate treatment effects, so the tree does not over-segment patients based on noise. However, the degree of this conservatism is controlled by the choice of complexity parameters, which involves a tradeoff between the granularity of personalization and the reliability of the resulting subgroups. Shallower trees produce fewer, larger subgroups that are easier to calibrate reliably but may collapse clinically distinct patient types into a single treatment rule. Deeper trees capture finer-grained heterogeneity and yield more targeted recommendations, but smaller subgroups are harder to calibrate, particularly when treatment exposure within a subgroup is limited.

A conservative approach, restricting tree depth to guarantee calibration, risks discarding clinically meaningful variation. Our framework offers an alternative: fit a deeper tree to capture the full heterogeneity signal, then use calibration to determine which subgroups are reliable enough to deploy. Subgroups that pass the calibration threshold can be acted upon; those that do not are flagged for caution rather than silently included. In this study, this approach identified four of five subgroups as deployment-ready while correctly flagging Group 1, the smallest subgroup with the fewest treated patients, as unreliable. The result is finer-grained personalization where the data support it, without sacrificing reliability where they do not.

#### 4.2.3 Calibration-guided implementation

Our framework enables selective deployment: rather than launching complex decision support systems across all patient segments simultaneously, healthcare systems can prioritize subgroups with low calibration error and clinically meaningful effect magnitude for initial rollout.

#### 4.3 Limitations

##### Study design

This study uses a single institution dataset (INSPIRE dataset, Seoul National University Hospital), limiting generalizability. Practice patterns, patient characteristics, and outcome rates differ across healthcare systems. The clinical scope is elective prostate procedures, so sex-based comparisons are not applicable in this case study. We assessed reliability across available clinical subgroups (age, BMI, ASA, diabetes), but this does not constitute a full equity audit across sociodemographic groups or multi-site settings. External validation of the method is required before broader clinical implementation.

##### Temporal recording challenges

Diagnosis timestamps appear to reflect time of recording rather than time of onset, as evidenced by many diagnoses falling outside the admission timeframe. Diagnosis features were therefore excluded to avoid potential bias. Future work should incorporate true diagnosis timing to enable their use as covariates.

##### Outcome measurement challenge

The opioid study used opioid medication count, a proxy for opioid usage that does not directly capture pain intensity, functional status, patient satisfaction, or adverse effects. Identical medication counts may reflect different pain control quality. Nonetheless, opioid medication count is a clinically meaningful endpoint, and the finding that neuraxial anesthesia reduces it by approximately 1.4 medications, is consistent with clinical expectations and warrants prospective validation. The primary contribution of this study is the methodological framework; the clinical finding serves as a proof-of-concept illustration.

##### Causal inference challenges

The prostate opioids study had a moderately imbalanced treatment distribution (28.7% treated vs. 71.3% control, 809 treated vs. 2,013 control). The overall sample (N=2,822) was sufficiently large for primary analyses, though treatment imbalance (28.7% vs. 71.3%) may have reduced power in subgroup comparisons (e.g., Group 1: 82 treated, 168 control), which may have limited heterogeneity discovery. In addition, imbalance of the age covariate (SMD = 0.236) warrants caution.

##### Effect-tree methodology

Effect-trees may be unstable with small subgroups and may under-represent continuous heterogeneity due to their discretization of continuous variables. The tree algorithm is sensitive to the choice of maximum depth and minimum leaf size, which should be carefully selected to address the business objectives of the analysis.

##### Missing covariates at implementation

The framework requires complete values for rule-defining covariates to assign a patient to an effect-tree subgroup. If one or more required covariates are unavailable at decision time, subgroup rules may not be applicable, limiting personalization for those patients.

##### Calibration assessment

Our calibration thresholds involve judgment about acceptable error magnitudes, which can change the determination of whether a subgroup is calibrated or not.

### 4.4 Future directions

#### Prospective trials

Randomized trials that compare algorithm-guided versus standard decision-making would provide the gold-standard for evidence, but face ethical and logistical challenges. External validation in independent cohorts across diverse institutions and populations is essential, particularly to assess the benefits of the approach as well as whether specific effect-tree rules generalize beyond the training population.

#### Methodological refinements

Methodological refinements could include effect-tree stability assessment via bootstrapping and uncertainty-aware tree algorithms accounting for CATE estimation error. In addition, distributional measures of within-group heterogeneity could be used to assess the certainty of the group ATE, and its applicability to individual patients. These measures could be used to further define implementation criteria.

#### Other clinical areas

The framework could extend to other perioperative interventions to systematically identify which treatments warrant personalized versus uniform strategies.

### 4.5 Conclusions

We demonstrated a framework for responsible treatment personalization from observational data, combining causal effect estimation, effect-tree subgroup discovery, and calibration assessment to determine whether heterogeneous effects warrant personalization versus population-level recommendations. Applied to neuraxial anesthesia in prostate procedure patients (N=2,822), the framework identified five subgroups using BMI, ASA status, and age. Four of five subgroups (91.1% of patients) showed large, well-calibrated effects (*−*1.29 to *−*1.59 opioid medications), supporting deployment. One subgroup (8.9%) showed poor calibration despite a predicted benefit, illustrating how the framework identifies unreliable heterogeneity. Together, these results reinforce that both effect magnitude and calibration assessment are central deployment criteria.

The contributions extend beyond perioperative medicine. Wherever heterogeneous treatment effects can be estimated from observational data, the same approach, effect-trees to generate interpretable rules, calibration to validate them, can guide decisions about which patient subgroups are ready for targeted clinical recommendations and which require further evidence. Method triangulation (five estimation methods) and sensitivity analyses assess robustness of causal claims. This moves causal machine learning from exploratory analysis toward validated decision support that can be deployed selectively and monitored over time.

## Data Availability

The INSPIRE dataset used in this study is publicly available as described by Lim et al.
Analytical code is available from the corresponding author upon reasonable request.

https://physionet.org/content/inspire/1.3/

## Disclosures

### Declaration of competing interests

All authors are associated with Atidia Health.

### Funding

This research was conducted as part of the authors’ employment at Atidia Health without any grant from funding agencies in the public, commercial, or not-for-profit sectors.

### Use of AI Tools

Generative AI tools (ChatGPT 5.3 Codex, Claude Sonnet 4.5, Claude Opus 4.6) were used to assist with language editing, literature search, code development and optimization, and visualization. All research questions, methodological decisions, interpretation of results, and scientific conclusions were determined by the authors. The study design, analytical approach, and validation of all outputs remained under full author control and responsibility. AI tools are not listed as authors and do not bear responsibility for the content.

### Ethics

This study used the INSPIRE dataset, a publicly available, fully de-identified perioperative dataset from Seoul National University Hospital, South Korea [14]. Ethical approval and patient consent for the original INSPIRE data collection were obtained by the data custodians. No additional ethics approval was required for secondary analysis of this publicly available de-identified dataset.

### Protocol and registration

No formal prospective protocol was prepared for this retrospective secondary analysis, and the study was not registered.

### Data availability

The INSPIRE dataset used in this study is publicly available as described by Lim et al. [14]. We analyzed the de-identified release and did not generate new patient-level data.

### Code availability

Analytical code is available from the corresponding author upon reasonable request.

### Patient and public involvement

Patients and members of the public were not involved in the design, conduct, reporting, or dissemination plans of this study.

## Notes

### Author Declarations

The inspire dataset is publicly available on physionet https://physionet.org/content/inspire

### Summary of Updates

There were no significant changes in manuscript content. An error in the citation of the INSPIRE dataset was fixed. Funding and competing interest statements were updated. Author details were updated.

