## Supplementary materials for "Trustworthy personalized treatment selection: causal effect-trees and calibration in perioperative medicine"

Mittelberg et al.

#### S1 Population Statistics and Study Characteristics

##### S1.1 INSPIRE Dataset Overview

**Dataset**: INSPIRE (INternational Surgical PerioperativE Registry)
**Institution**: Seoul National University Hospital
**Description**: Comprehensive perioperative dataset containing electronic health record data from surgical patients

**Dataset Statistics**

- **Total Admissions**: 126,673
- **Total Unique Patients**: 99,886
- **Total Operations**: 130,960
- **Total Diagnosis Records**: 2,464,620
- **Total Laboratory Records**: 19,503,335
- **Total Medication Records**: 9,885,572

##### S1.2 Neuraxial Anesthesia and Post-Operative Opioid Use in Prostate Procedures

**Study Population**

**Total Sample Size**: N = 2,822
**Treatment Groups**:

- Treated (neuraxial anesthesia): 809 (28.7%)
- Control (general anesthesia): 2,013 (71.3%)

**Outcome**: Post-operative opioid medication count within 48 hours after surgery (N02A ATC codes)

#### Participant Selection Flow (Supplementary Table S1)

**All operations in raw operations.csv (N=130,960)**

↓ Operation-level aggregation to one row per `hadm_id`, removed $n = 4,287$

**Admission-level base (N=126,673)**

↓ Exclude non-prostate procedures, $n = 123,835$

**Prostate procedures (N=2,838)**

↓ Apply age restriction [18, 90], removed $n = 0$

**Age-eligible prostate procedures (N=2,838)**

↓ Exclude female sex, $n = 1$

**Male prostate procedures (N=2,837)**

↓ Exclude emergency procedures (`emop=1`), $n = 15$

**Non-emergency male prostate procedures (N=2,822)**

↓ Apply anesthesia type restriction (General/Neuraxial), removed $n = 0$

**Post-population-filter cohort (N=2,822)**

↓ Exclude missing outcome (48h opioid count), removed $n = 0$

**Final analytic cohort with outcome observed (N=2,822)**

Figure S1: Participant flow diagram for cohort derivation, including outcome completeness (0 missing outcomes). Exact stepwise counts are reported in Supplementary Table S1.

#### Inclusion Criteria

- Prostate procedures (identified by prostate procedure indicator, ICD-10-PCS codes 0VT0, 0VB0)
- Age 18–90 years
- Male patients only
- General or neuraxial anesthesia types only

#### Exclusion Criteria

- Emergency procedures

#### Adjustment Covariates

Four baseline covariates were used for both confounding adjustment and heterogeneity estimation:

- BMI (body mass index)
- ASA (ASA physical status classification)
- Age
- Diabetes Indicator (medication and laboratory based)

#### Baseline Characteristics (Supplementary Table S2)

| Characteristic | Treated (N=809) | Control (N=2,013) | SMD | Balance |
|---|---|---|---|---|
| Bmi | $24.3 \pm 3.0$ | $24.7 \pm 3.7$ | $-0.131$ | Acceptable ($< 0.25$) |
| Asa | $1.8 \pm 0.5$ | $1.8 \pm 0.5$ | $-0.061$ | Good ($< 0.1$) |
| Age | $69.6 \pm 7.2$ | $67.9 \pm 7.4$ | 0.236 | Acceptable ($< 0.25$) |
| Diabetes Indicator (%) | 7.7% | 7.7% | $-0.003$ | Good ($< 0.1$) |
| Opioid Use 48H | $0.52 \pm 1.04$ | $1.84 \pm 1.65$ | — | — |

Table S1: Baseline characteristics by treatment group

Values are mean ± SD, or % for binary indicators. SMD: standardized mean difference; Balance: Good $< 0.1$, Acceptable $< 0.25$.

**Pre-Imputation Variable Missingness (Supplementary Table S3)**

| Variable | Missing, n | Missing, % |
|---|---|---|
| Treatment (neuraxial vs general anesthesia) | 0 | 0.0000 |
| Outcome (48h post-operative opioid medication count) | 0 | 0.0000 |
| Age | 0 | 0.0000 |
| BMI | 15 | 0.5315 |
| ASA status | 17 | 0.6024 |
| Diabetes indicator | 0 | 0.0000 |
| Prior opioid use indicator | 0 | 0.0000 |

Table S2: Pre-imputation missingness for variables used in the analytic model (cohort after population filters, N=2,822)

### S2 Additional Detailed Results for Neuraxial Anesthesia Study (Prostate Procedures)

This section provides detailed results from the neuraxial versus general anesthesia case study in prostate procedures that were condensed in the main text for brevity.

#### Calibration by CATE Quintiles

To evaluate model reliability for personalized treatment decisions, we assessed calibration across quintiles of predicted treatment effects. Patients were binned into five groups based on their individualized treatment effect predictions:

| Quintile | Predicted | Observed | Error | $N_{treated}$ | $N_{control}$ | Ratio |
|---|---|---|---|---|---|---|
| Q1 (Largest Benefit) | −1.602 | −1.591 | 0.011 | 177 | 389 | 0.46:1 |
| Q2 | −1.482 | −1.598 | 0.116 | 140 | 423 | 0.33:1 |
| Q3 | −1.367 | −1.217 | 0.150 | 167 | 397 | 0.42:1 |
| Q4 | −1.284 | −1.256 | 0.028 | 172 | 407 | 0.42:1 |
| Q5 (Smallest Benefit) | −1.174 | −0.918 | 0.256 | 153 | 397 | 0.39:1 |

Table S3: Detailed Calibration Assessment by Predicted Treatment Effect Quintiles

**Calibration Quality** Mean absolute calibration error was 0.112 (6.9% of outcome standard deviation = 1.62), indicating good calibration overall. Calibration errors ranged from 0.011 (Q1, good) to 0.256 (Q5, moderate). The treatment ratios across quintiles indicate adequate positivity across the range of predicted effects, although the treated group is consistently smaller (28.7% overall).

**Ranking Reliability** The model demonstrated strong ranking reliability: predicted and observed effects showed strong correlation ($r = 0.937$). A non-monotonic relationship was detected between Q1 and Q2 (observed effect slightly larger for Q2 than Q1), suggesting some noise in the extreme quantiles.

**Clinical Interpretation** The calibration results support the model's ability to reliably rank patients by expected benefit magnitude. The largest calibration error occurs in Q5 (smallest predicted benefit), consistent with the effect tree finding that low-BMI, low-ASA patients (Group 1) have less reliable effect estimates.

#### Detailed Effect Tree Subgroup Characterization

Beyond the summary provided in the main text, complete characterization of effect tree subgroups reveals:

**Group 1** (BMI $\leq$ 22.87, ASA $\leq$ 1.50; N=250) Predicted effect −1.10 (observed −0.66; calibration error 0.44). This is the smallest subgroup (8.9%), containing young, lower BMI, healthy patients (mean age 67.7, mean BMI 21.2, ASA 1.0, diabetes 2.0%). Treatment balance: 82 treated vs 168 control. The high calibration error (predicted effect overestimates observed benefit by 68%) suggests caution in deploying targeted recommendations for this subgroup. **Recommendation: Consider carefully.**

**Group 2** (BMI $\leq$ 22.87, ASA > 1.50; N=592) Predicted effect $-1.29$ (observed $-1.35$; calibration error 0.05). This subgroup contains lower BMI patients with higher comorbidity burden (mean age 70.0, mean BMI 21.1, mean ASA 2.1, diabetes 10.0%). Treatment balance: 176 treated vs 416 control. Good calibration with the observed effect slightly exceeding the prediction. **Recommendation: Implement.**

**Group 3** (BMI > 22.87, age $\leq$ 72.50, BMI $\leq$ 26.28; N=993) Predicted effect $-1.34$ (observed $-1.27$; calibration error 0.07). The largest subgroup (35.2%), containing normal-weight younger patients (mean age 64.8, mean BMI 24.8, mean ASA 1.8, diabetes 7.5%). Treatment balance: 268 treated vs 725 control. **Recommendation: Implement.**

**Group 4** (BMI > 22.87, age $\leq$ 72.50, BMI > 26.28; N=468) Predicted effect $-1.49$ (observed $-1.47$; calibration error 0.02). Contains overweight/obese younger patients (mean age 64.4, mean BMI 29.0, mean ASA 1.8, diabetes 8.5%). Treatment balance: 111 treated vs 357 control. The best-calibrated subgroup. **Recommendation: Implement.**

**Group 5** (BMI > 22.87, age > 72.50; N=519) Predicted effect $-1.59$ (observed $-1.56$; calibration error 0.04). Contains older patients with higher BMI (mean age 77.0, mean BMI 25.8, mean ASA 1.9, diabetes 7.7%). Treatment balance: 172 treated vs 347 control. Shows the largest predicted and observed benefit. **Recommendation: Implement.**

##### Summary

The effect tree identified five clinically interpretable subgroups using BMI, ASA status, and age. The primary split on BMI (threshold 22.87) separates lower BMI from higher BMI patients, with secondary splits on ASA (for lower BMI patients) and age (for higher BMI patients). All five subgroups show predicted benefit, with effects ranging from $-1.10$ to $-1.59$ opioid medications. Four of five subgroups (91.1% of patients) demonstrate good calibration (errors 0.02–0.07), supporting selective deployment of treatment recommendations. The poorly calibrated subgroup (Group 1, N=250, calibration error 0.44) consists of lower BMI, healthy patients where the model overestimates the benefit magnitude.

#### S3 Detailed Feature-Based Subgroup Analyses

##### Method

For each adjustment covariate, we tested whether individual CATE estimates varied systematically with covariate value. For continuous covariates (BMI, age, ASA status), we computed the Spearman rank correlation between the covariate and individual CATE estimates; a negative correlation indicates that higher covariate values are associated with larger (more negative) treatment effects. For binary covariates (diabetes), we used the Mann-Whitney U test to compare CATE distributions between groups. The covariate *prior opioid use* was excluded from the analysis as it was constant (zero for all patients) in this dataset.

##### Results

All four tested covariates showed statistically significant effect modification (Supplementary Table S5). The direction and strength of associations are consistent with the effect tree structure: Group 5 (older patients with higher BMI) showed the largest benefit ($-1.59$ medications) while Group 1 (lower BMI, low ASA) showed the smallest ($-1.10$ medications).

| Covariate | Type | Test | Statistic | p-value | Direction |
|---|---|---|---|---|---|
| BMI | Continuous | Spearman $r$ | $-0.588$ | $< 0.001$ | Higher BMI → larger benefit |
| Age | Continuous | Spearman $r$ | $-0.385$ | $< 0.001$ | Older age → larger benefit |
| ASA status | Continuous | Spearman $r$ | $-0.229$ | $< 0.001$ | Higher ASA → larger benefit |
| Diabetes | Binary | Mann-Whitney $U$ | 317,349 | 0.004 | Diabetic → larger benefit |

Table S4: Feature-based heterogeneity tests. Individual CATE estimates from the causal forest DML model were correlated with each covariate to assess effect modification.

Supplementary Figure S2 visualizes these relationships, showing individual CATE estimates plotted against each covariate with the test statistic annotated on each panel.

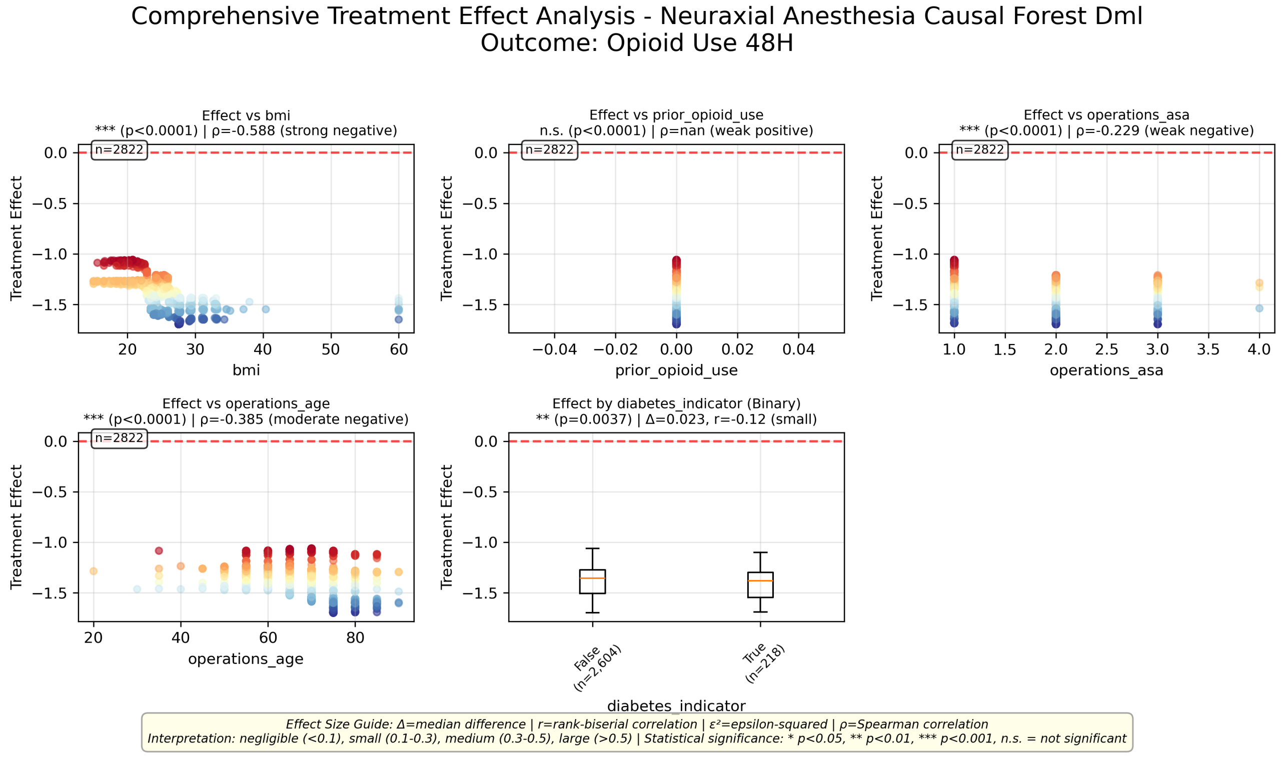


Figure S2: Feature-based heterogeneity analysis. Each panel plots individual CATE estimates (causal forest DML) against a covariate. For continuous and ordinal covariates (BMI, ASA status, age), Spearman $r$ and its significance are shown; a negative $r$ indicates higher covariate values are associated with larger (more negative) treatment effects. For the binary covariate (diabetes), a boxplot compares CATE distributions between groups with the median difference ($\Delta = \text{median}_{\text{non-diabetic}} - \text{median}_{\text{diabetic}} = +0.023$, i.e., diabetic patients have slightly larger benefit) and rank-biserial correlation ($|r| = 0.12$, negligible effect size). Prior opioid use was constant across patients (all zero) and is shown for completeness.

##### Convergence with effect tree and causal forest feature importance

These results provide triangulating evidence for the effect modifiers identified by the effect tree. Three independent methods — effect tree node splits, causal forest feature importance, and the Spearman correlations above — converge on the same three variables in the same rank order: BMI (feature importance 0.676, $|r| = 0.588$) is the strongest modifier, followed by age (0.206, $|r| = 0.385$) and ASA status (0.087, $|r| = 0.229$). This convergence increases confidence that the effect tree splits reflect genuine causal heterogeneity rather than artifacts of the tree-fitting algorithm.

Diabetes was statistically significant in the feature-based analysis ($p = 0.004$) but was not selected by the effect tree and has negligible causal forest feature importance. The median CATE difference between diabetic and non-diabetic patients is only 0.023 opioid medications ($|r| = 0.12$, a negligible effect size).

### S4 Case-Study Run Configuration

This section summarizes the pre-specified run settings used for the prostate neuraxial-anesthesia case study.

| **Component** | **Configuration used** |
|---|---|
| Global reproducibility | Random seed: 42 |
| Propensity model | XGBoost; 100 trees; max depth 4; learning rate 0.1; 5-fold CV |
| CausalForestDML (primary HTE model) | Random-forest base model; 400 trees; minimum samples per leaf 50; max sample fraction 0.5; honest splitting enabled; 5-fold cross-fitting; inference enabled; CI alpha 0.05 |
| Effect-tree interpreter | SingleTreeCateInterpreter; max depth 3; minimum samples per leaf 200 |
| IPW | trimming threshold 0.10 |
| Doubly robust (DR) | random-forest nuisance/final models |
| Meta-learners | S-learner (random forest), X-learner (random forest) |
| Sensitivity analysis | E-value and placebo tests |
| Calibration thresholds | Good $< 0.10$ SD; Moderate $< 0.25$ SD |
| Implementation effect thresholds | Minimal effect: 0.2 opioid medications; Meaningful/Substantial effect: 1.0 opioid medication |

Table S5: Estimator and analysis settings used in the prostate case study
